# Chronic kidney disease in Low- and Middle- Income Countries: Protocol for a systematic review of diagnostic and prognostic models

**DOI:** 10.1101/2021.04.24.21256041

**Authors:** Edson J Ascencio, Diego J Aparcana-Granda, Rodrigo M Carrillo-Larco

**Author notes:** **Corresponding author** Rodrigo M Carrillo-Larco, MD, Department of Epidemiology and Biostatistics, School of Public Health, Imperial College London, London W2 1PG, UK. **Competing interests:** The authors declare no conflict of interests.

## Abstract

**Background:** Chronic Kidney Disease (CKD) is a highly prevalent condition with a large disease burden globally. In low- and middle-income countries (LMIC) the CKD screening challenges the health system. This systematic and comprehensive search of all CKD diagnostic and prognostic models in LMIC will inform screening strategies in LMIC following a risk-based approach.

**Objective:** To summarize all multivariate diagnostic and prognostic models for CKD in adults in LMIC.

**Methods:** Systematic review. Without date or language restrictions we will search Embase, Medline, Global Health (these three through Ovid), SCOPUS and Web of Science. We seek multivariable diagnostic or prognostic models which included a random sample of the general population. We will screen titles and abstracts; we will then study the selected reports. Both phases will be done by two reviewers independently. Data extraction will be performed by two researchers independently using a pre-specified Excel form (CHARMS model). We will evaluate the risk of bias with the PROBAST tool.

**Conclusion:** This systematic review will provide the most comprehensive list and critical appraisal of diagnostic and prognostic models for CKD available for the general population in LMIC. This evidence could inform policies and interventions to improve CKD screening in LMIC following a risk-based approach, maximizing limited resources and reaching populations with limited access to CKD screening tests. This systematic review will also reveal methodological limitations and research needs to improve CKD diagnostic and prognostic models in LMIC.

## INTRODUCTION

Chronic kidney disease (CKD) is a highly prevalent condition that contributes to a large part of disease burden globally. Between 1990 and 2017, the health metrics of CKD showed a bleak profile: mortality rate, incidence and kidney transplantation rate increased by 2.8%, 29.3% and 34.4%, respectively.^1^ CKD led to 1.2 million deaths in 2017 and in the best-case scenario, mortality is projected to increase to 2.2 million deaths^2^ and become the 5th cause of years of life lost (YLL) by 2040.^3^ Currently, 2.5 million of patients receive kidney transplantation therapy and it is projected to increase to 5.4 million by 2030.^1^ CKD also reveals disparities between low- and middle-income countries (LMIC) and high income countries (HIC); for example, the age-standardised disability-adjusted life-year (DALY) rate due to CKD was the highest in LMIC between 1990-2017.^4^ In LMIC, that remain as resource-constrained settings, there is a need for optimization of the CKD screening strategies which usually challenge the health system.^5^

Risk equations or risk scores are a cost-effective alternative for CKD screening.^6^ These equations are less invasive and accepted by the general population;^7^ also, they require less resources like laboratory tests.^8^ Many scores were developed in high-income countries,^9-11^ and they may not be used in LMIC because their accuracy is better where they have been developed.^12^ Current strategies for CKD screening suggest studying people with risk factors (e.g. diabetes, hypertension).^13-15^ These recommendations rely on studies where albuminuria and proteinuria were used as screening tools for identifying CKD patients.^16^ Nevertheless, a systematic review found that using risk scores allows screening of a larger population and therefore can be useful for detecting more CKD cases.^6^

To date, there are no systematic reviews of diagnostic or prognostic models for CKD with a focus on LMIC.^17, 18^ This limits our knowledge of what tools we have to enhance CKD screening in LMIC; similarly, this dearth of evidence prevents from planning future research to overcome the limitations of available models. This will be the first systematic review to fill these knowledge gaps in LMIC to improve and complement the CKD screening programmes in LMIC.

## METHODS

### Objective

To synthesise CKD diagnostic and prognostic models for the adult population of LMIC.

### Study design

This systematic review and meta-analysis will be conducted following the preferred reporting items for systematic reviews and meta-analyses (PRISMA) 2020 guidelines.^19^ We will also adhere to the recommendations for systematic reviews of diagnostic and prognostic models following the CHARMS guidelines (Table 1)^20^ and the PROBAST tool to assess risk of bias.^21^

**Table 1:** CHARMS criteria to define research question and strategy.

| Concept | Criteria |
| --- | --- |
| Prognostic or diagnostic? | Both - this review will focus on diagnostic and prognostic risk scores for chronic kidney disease (CKD). |
| Scope | Diagnostic/prognostic models to inform physicians, researchers and the general population whether they are likely to have CKD (i.e. diagnostic) or will be likely to have CKD (i.e. prognostic). |
| Type of prediction modelling studies | • Diagnostic/prognostic models with external validation<br>• Diagnostic/prognostic models without external validation<br>• Diagnostic/prognostic models validation |
| Target population to whom the prediction model applies | General and hospital-based adult population in Low- and Middle-income Countries (LMIC), no age or gender restrictions. |
| Outcome to be predicted | CKD (diagnostic or prognostic). |
| Time span of prediction | Any, prognostic models will not be included/excluded based on the prediction time span. |
| Intended moment of using the model | Diagnostic/prognostic models to be used in asymptomatic adults of LMIC to ascertain current CKD status (i.e diagnostic) or future risk of developing CKD (i.e. prognostic). These models could be used for screening, treatment allocation in primary prevention or research purposes. |

Based on the CHARMS checklist.

### Eligibility criteria

#### Participants/population

We will include the general adult population (18 years and above) of LMIC with no gender restrictions. Studies following a population-based random sampling approach will be included. We will only include populations from LMIC according to The World Bank.^22^ Conversely, studies with a study population of only patients (e.g., people with hypertension) or high-risk individuals (e.g., smokers) will be excluded. We will exclude studies with LMIC populations outside a LMIC.

#### Intervention, exposure

None (this review is looking at CKD diagnostic and prognostic models in LMIC).

#### Comparator, control

None (this review is looking at CKD diagnostic and prognostic models in LMIC).

#### Outcome

Diagnostic and prognostic models for CKD. The CKD diagnosis should have been based on a laboratory or imaging test including: urine albumin-creatinine ratio, urine protein-creatinine ratio, albumin excretion ratio, urine sediment, kidney images, kidney biopsy or the estimated glomerular filtration rate (eGFR). In other words, research in which CKD diagnosis was based on self-reported information only will not be considered. However, if a study combined both self-reported information and a laboratory or imaging tests, this will be included.

#### Types of studies

Studies with an observational design will be included, which encompasses cross-sectional (for diagnostic models) and prospective longitudinal studies (for prognostic models). If we retrieve any systematic review on this subject, we will revise its reference list to identify relevant original sources.

### Literature Search and Data collation

The search will be conducted in five search engines: Embase, Medline, Global Health (these three through Ovid), SCOPUS and Web of Science. No date or language restrictions will be set. The complete search strategy can be found in Supplementary Material.

Titles and abstracts will be screened by two researchers independently (DJA-G and EJA), looking for studies that meet the selection criteria above detailed. Full-text reports of the selected publications will be studied by two researchers independently (DJA-G and EJA). Discrepancies at any stage will be solved by consensus or by a third party (RMC-L).

During the full-text phase, if there are any original reports in which the population, methodology or results are not clear enough to assess the inclusion/exclusion criteria, we will contact the corresponding author by email. We will wait for two weeks, if we receive no answer and cannot solve our doubts through other means, this report will be excluded based on the lack of clarity to assess inclusion/exclusion criteria.

We will record the reasons for exclusion in the full-text phase and summarize the number of included/excluded reports following the PRISMA flow diagram.

### Data extraction

We will develop a data extraction form following the CHARMS recommendations.^20^ Data extraction will be conducted by two researchers independently; discrepancies will be solved by consensus or by a third party (RMC-L).

### Risk of bias of individual studies

The risk of bias assessment of individual reports will be conducted using the Prediction model Risk Of Bias ASsessment Tool (PROBAST) tool.^21^

### Statistical Analysis

A qualitative synthesis is planned, whereby we will narratively synthesise the findings from the selected studies. We will summarize the key elements from each report such as study design, study population and characteristics of the study population. Also, we will summarize the key features of the risk scores as provided by each report, including discrimination, calibration, sensitivity, specificity, and predictive values. A quantitative synthesis will be carried out if the included studies are found to be sufficiently homogenous and we have at least four original reports.

### Ethics

This review did not directly include human subjects. We considered this work as ‘low risk’ and did not request approval by an Ethics Committee. Results and opinions included in this protocol, and those included in the final report, are the author’s alone and do not represent those of the institutions to which they belong.

## CONCLUSIONS

This systematic review will provide a comprehensive list of diagnostic and prognostic models for CKD for people in LMIC, along with their accuracy metrics. Currently, information lacks in LMIC where diagnostic and prognostic models could inform CKD screening strategies. Similarly, this work will elucidate the limitations of available diagnostic and prognostic models for CKD in LMIC, so that future research can be planned accordingly to overcome these caveats and deliver robust models to advance CKD screening strategies in LMIC.

## Data Availability

Supplementary materials will be presented with the main protocol.

## SUPPLEMENTARY MATERIAL

### OVID (Medline, Embase, Global Health)

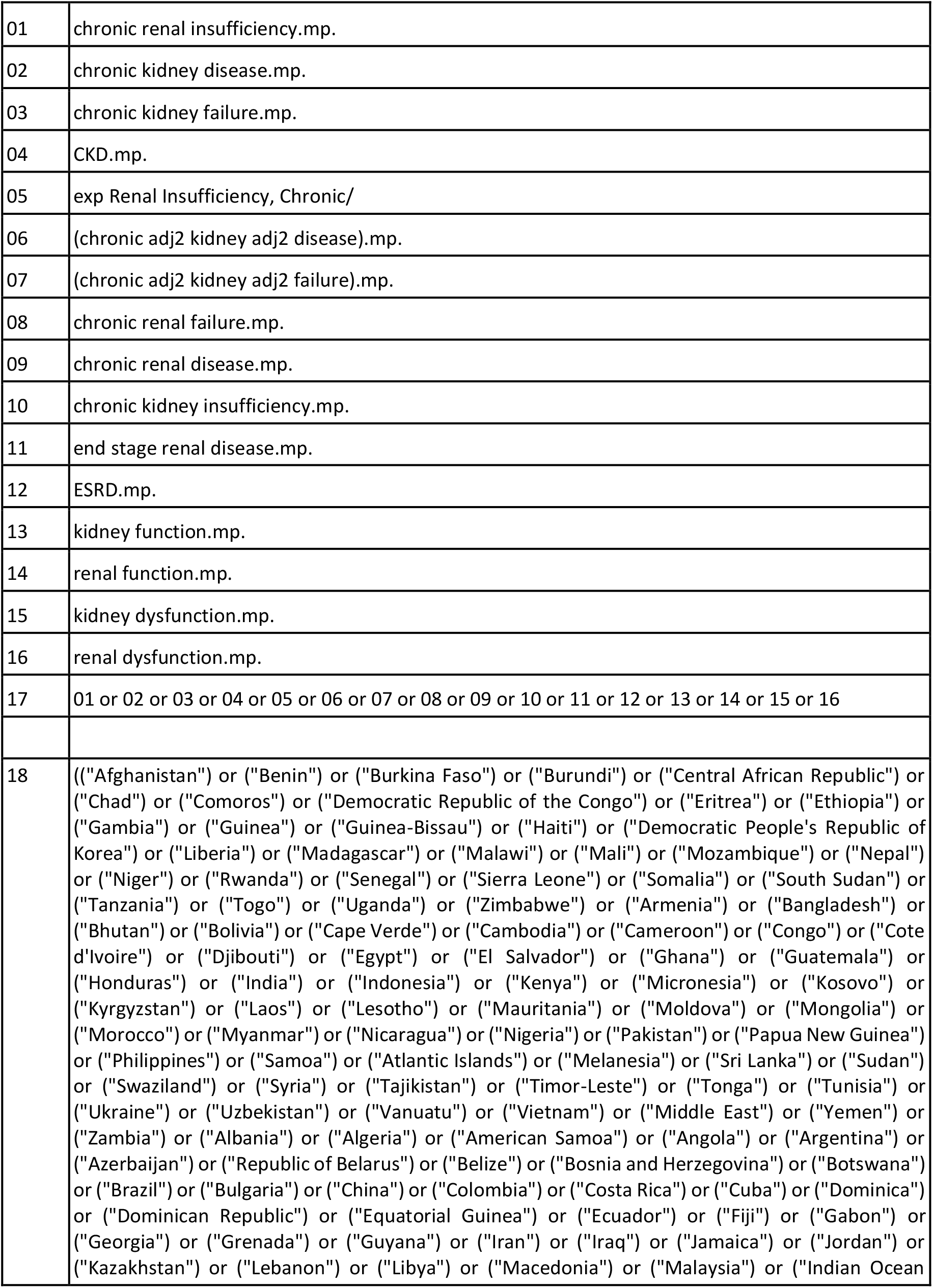
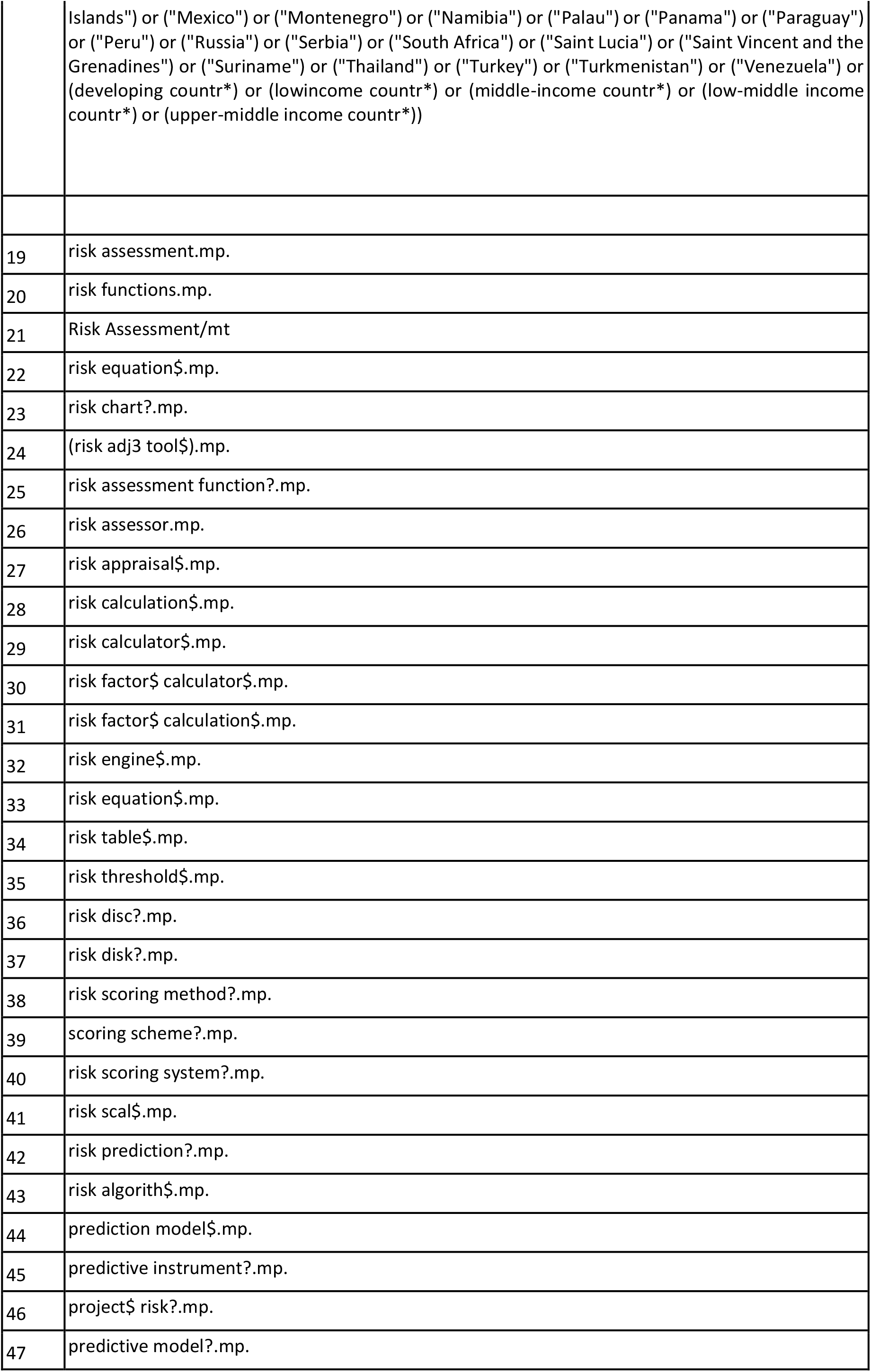
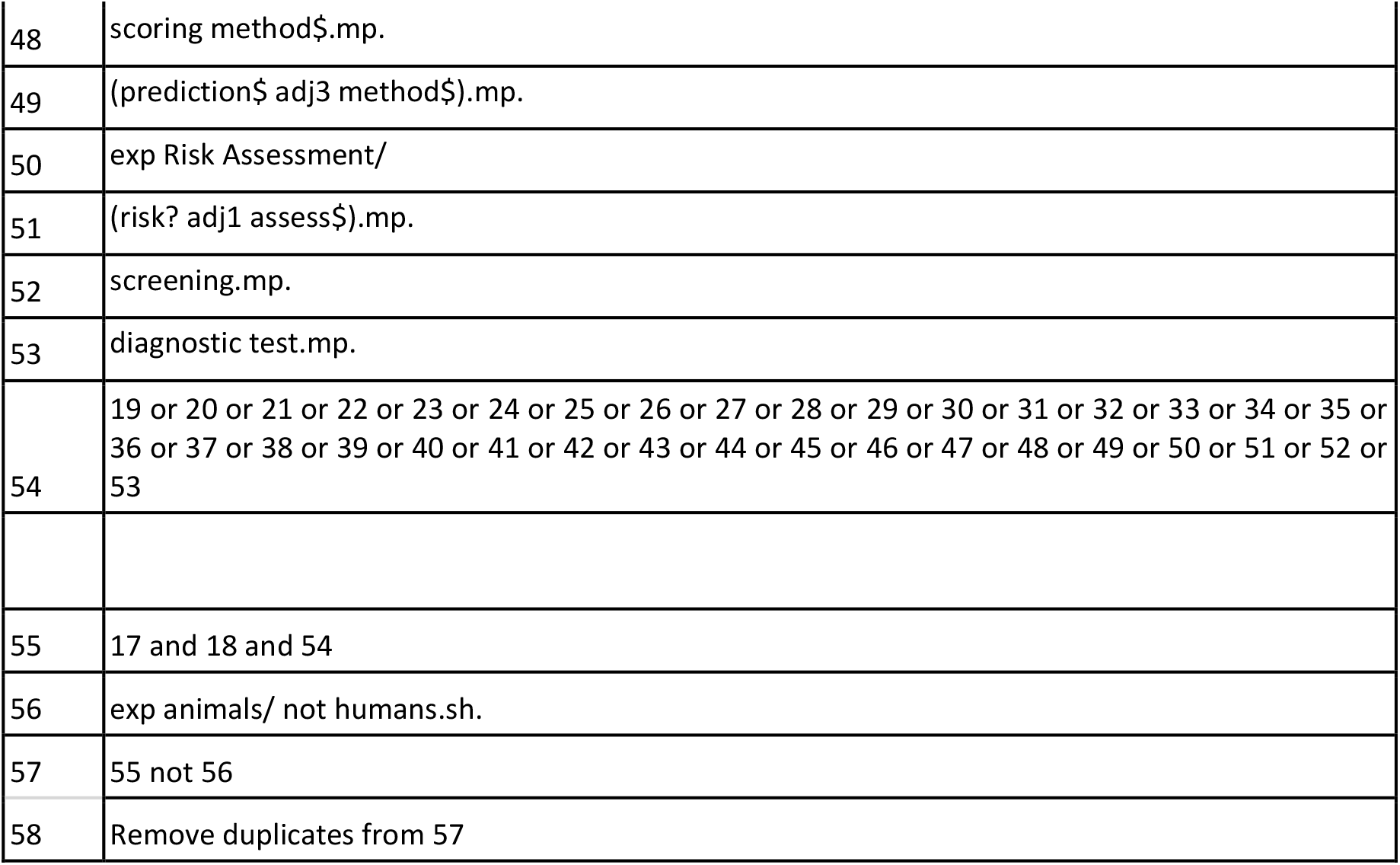

### SCOPUS

((TITLE-ABS-KEY(“Afghanistan”) OR TITLE-ABS-KEY(“Benin”) OR TITLE-ABS-KEY(“Burkina Faso”) OR TITLE-ABS-KEY(“Burundi”) OR TITLE-ABS-KEY(“Central African Republic”) OR TITLE-ABS-KEY(“Chad”) OR TITLE-ABS-KEY(“Comoros”) OR TITLE-ABS-KEY(“Democratic Republic of the Congo”) OR TITLE-ABS-KEY(“Eritrea”) OR TITLE-ABS-KEY(“Ethiopia”) OR TITLE-ABS-KEY(“Gambia”) OR TITLE-ABS-KEY(“Guinea”) OR TITLE-ABS-KEY(“Guinea-Bissau”) OR TITLE-ABS-KEY(“Haiti”) OR TITLE-ABS-KEY(“Democratic People’s Republic of Korea”) OR TITLE-ABS-KEY(“Liberia”) OR TITLE-ABS-KEY(“Madagascar”) OR TITLE-ABS-KEY(“Malawi”) OR TITLE-ABS-KEY(“Mali”) OR TITLE-ABS-KEY(“Mozambique”) OR TITLE-ABS-KEY(“Nepal”) OR TITLE-ABS-KEY(“Niger”) OR TITLE-ABS-KEY(“Rwanda”) OR TITLE-ABS-KEY(“Senegal”) OR TITLE-ABS-KEY(“Sierra Leone”) OR TITLE-ABS-KEY(“Somalia”) OR TITLE-ABS-KEY(“South Sudan”) OR TITLE-ABS-KEY(“Tanzania”) OR TITLE-ABS-KEY(“Togo”) OR TITLE-ABS-KEY(“Uganda”) OR TITLE-ABS-KEY(“Zimbabwe”) OR TITLE-ABS-KEY(“Armenia”) OR TITLE-ABS-KEY(“Bangladesh”) OR TITLE-ABS-KEY(“Bhutan”) OR TITLE-ABS-KEY(“Bolivia”) OR TITLE-ABS-KEY(“Cape Verde”) OR TITLE-ABS-KEY(“Cambodia”) OR TITLE-ABS-KEY(“Cameroon”) OR TITLE-ABS-KEY(“Congo”) OR TITLE-ABS-KEY(“Cote d’Ivoire”) OR TITLE-ABS-KEY(“Djibouti”) OR TITLE-ABS-KEY(“Bolivia”) OR TITLE-ABS-KEY(“Cape Verde”) OR TITLE-ABS-KEY(“Cambodia”) OR TITLE-ABS-KEY(“Cameroon”) OR TITLE-ABS-KEY(“Congo”) OR TITLE-ABS-KEY(“Cote d’Ivoire”) OR TITLE-ABS-KEY(“Djibouti”) OR TITLE-ABS-KEY(“Egypt”) OR TITLE-ABS-KEY(“El Salvador”) OR TITLE-ABS-KEY(“Ghana”) OR TITLE-ABS-KEY(“Guatemala”) OR TITLE-ABS-KEY(“Honduras”) OR TITLE-ABS-KEY(“India”) OR TITLE-ABS-KEY(“Indonesia”) OR TITLE-ABS-KEY(“Kenya”) OR TITLE-ABS-KEY(“Micronesia”) OR TITLE-ABS-KEY(“Kosovo”) OR TITLE-ABS-KEY(“Kyrgyzstan”) OR TITLE-ABS-KEY(“Laos”) OR TITLE-ABS-KEY(“Lesotho”) OR TITLE-ABS-KEY(“Mauritania”) OR TITLE-ABS-KEY(“Moldova”) OR TITLE-ABS-KEY(“Mongolia”) OR TITLE-ABS-KEY(“Morocco”) OR TITLE-ABS-KEY(“Myanmar”) OR TITLE-ABS-KEY(“Nicaragua”) OR TITLE-ABS-KEY(“Nigeria”) OR TITLE-ABS-KEY(“Pakistan”) OR TITLE-ABS-KEY(“Papua New Guinea”) OR TITLE-ABS-KEY(“Philippines”) OR TITLE-ABS-KEY(“Samoa”) OR TITLE-ABS-KEY(“Atlantic Islands”) OR TITLE-ABS-KEY(“Melanesia”) OR TITLE-ABS-KEY(“Sri Lanka”) OR TITLE-ABS-KEY(“Sudan”) OR TITLE-ABS-KEY(“Swaziland”) OR TITLE-ABS-KEY(“Syria”) OR TITLE-ABS-KEY(“Tajikistan”) OR TITLE-ABS-KEY(“Timor-Leste”) OR TITLE-ABS-KEY(“Tonga”) OR TITLE-ABS-KEY(“Tunisia”) OR TITLE-ABS-KEY(“Ukraine”) OR TITLE-ABS-KEY(“Uzbekistan”) OR TITLE-ABS-KEY(“Vanuatu”) OR TITLE-ABS-KEY(“Vietnam”) OR TITLE-ABS-KEY(“Middle East”) OR TITLE-ABS-KEY(“Yemen”) OR TITLE-ABS-KEY(“Zambia”) OR TITLE-ABS-KEY(“Albania”) OR TITLE-ABS-KEY(“Algeria”) OR TITLE-ABS-KEY(“American Samoa”) OR TITLE-ABS-KEY(“Angola”) OR TITLE-ABS-KEY(“Argentina”) OR TITLE-ABS-KEY(“Azerbaijan”) OR TITLE-ABS-KEY(“Republic of Belarus”) OR TITLE-ABS-KEY(“Belize”) OR TITLE-ABS-KEY(“Bosnia and Herzegovina”) OR TITLE-ABS-KEY(“Botswana”) OR TITLE-ABS-KEY(“Brazil”) OR TITLE-ABS-KEY(“Bulgaria”) OR TITLE-ABS-KEY(“China”) OR TITLE-ABS-KEY(“Colombia”) OR TITLE-ABS-KEY(“Costa Rica”) OR TITLE-ABS-KEY(“Cuba”) OR TITLE-ABS-KEY(“Dominica”) OR TITLE-ABS-KEY(“Dominican Republic”) OR TITLE-ABS-KEY(“Equatorial Guinea”) OR TITLE-ABS-KEY(“Ecuador”) OR TITLE-ABS-KEY(“Fiji”) OR TITLE-ABS-KEY(“Gabon”) OR TITLE-ABS-KEY(“Georgia”) OR TITLE-ABS-KEY(“Grenada”) OR TITLE-ABS-KEY(“Guyana”) OR TITLE-ABS-KEY(“Iran”) OR TITLE-ABS-KEY(“Iraq”) OR TITLE-ABS-KEY(“Jamaica”) OR TITLE-ABS-KEY(“Jordan”) OR TITLE-ABS-KEY(“Kazakhstan”) OR TITLE-ABS-KEY(“Lebanon”) OR TITLE-ABS-KEY(“Libya”) OR TITLE-ABS-KEY(“Macedonia (Republic)”) OR TITLE-ABS-KEY(“Malaysia”) OR TITLE-ABS-KEY(“Indian Ocean Islands”) OR TITLE-ABS-KEY(“Mexico”) OR TITLE-ABS-KEY(“Montenegro”) OR TITLE-ABS-KEY(“Namibia”) OR TITLE-ABS-KEY(“Palau”) OR TITLE-ABS-KEY(“Panama”) OR TITLE-ABS-KEY(“Paraguay”) OR TITLE-ABS-KEY(“Peru”) OR TITLE-ABS-KEY(“Russia”) OR TITLE-ABS-KEY(“Serbia”) OR TITLE-ABS-KEY(“South Africa”) OR TITLE-ABS-KEY(“SaintLucia”) OR TITLE-ABS-KEY(“Saint Vincent and the Grenadines”) OR TITLE-ABS-KEY(“Suriname”) OR TITLE-ABS-KEY(“Thailand”) OR TITLE-ABS-KEY(“Turkey”) OR TITLE-ABS-KEY(“Turkmenistan”) OR TITLE-ABS-KEY(“Venezuela”) OR TITLE-ABS-KEY(developing countr*) OR TITLE-ABS-KEY(lowincome countr*) OR TITLE-ABS-KEY(middle-income countr*) OR TITLE-ABS-KEY(low-middle income countr*) OR TITLE-ABS-KEY(upper-middle income countr*) OR TITLE-ABS-KEY(“low resource”) OR TITLE-ABS-KEY (“under-resourced”) OR TITLE-ABS-KEY(“resource poor”) OR TITLE-ABS-KEY(“under-developed”) OR TITLE-ABS-KEY(“underdeveloped”) OR TITLE-ABS-KEY(“developing world”) OR TITLE-ABS-KEY(“third world”) OR TITLE-ABS-KEY(lmic) OR TITLE-ABS-KEY(low AND middle AND income)) AND (TITLE-ABS-KEY(Risk Assessment) OR TITLE-ABS-KEY(risk? adj1 assess*) OR TITLE-ABS-KEY(risk function) OR TITLE-ABS-KEY(Risk Assessment) OR TITLE-ABS-KEY(risk functions) OR TITLE-ABS-KEY(risk equation*) OR TITLE-ABS-KEY(risk chart?) OR TITLE-ABS-KEY(risk adj3 tool*) OR TITLE-ABS-KEY(risk assessment function?) OR TITLE-ABS-KEY(risk assessor) OR TITLE-ABS-KEY(risk appraisal*) OR TITLE-ABS-KEY(risk calculation*) OR TITLE-ABS-KEY(risk calculator*) OR TITLE-ABS-KEY(risk factor*calculator*) OR TITLE-ABS-KEY(risk factor*calculation*) OR TITLE-ABS-KEY(risk engine*) OR TITLE-ABS-KEY(risk equation*) OR TITLE-ABS-KEY(risk table*) OR TITLE-ABS-KEY(risk threshold*) OR TITLE-ABS-KEY(risk disc?) OR TITLE-ABS-KEY(risk disk?) OR TITLE-ABS-KEY(risk scoring method?) OR TITLE-ABS-KEY(scoring scheme?) OR TITLE-ABS-KEY(risk scoring system?) OR TITLE-ABS-KEY(risk prediction?) OR TITLE-ABS-KEY(risk algorith*) OR TITLE-ABS-KEY(prediction model*) OR TITLE-ABS-KEY(predictive instrument?) OR TITLE-ABS-KEY(project*risk?) OR TITLE-ABS-KEY(predictive model?) OR TITLE-ABS-KEY(scoring method*) OR TITLE-ABS-KEY(prediction*adj3 method*) OR TITLE-ABS-KEY(screening) OR TITLE-ABS-KEY(risk scal*) OR TITLE-ABS-KEY(diagnostic test)) AND (TITLE-ABS-KEY(chronic renal insufficiency) OR TITLE-ABS-KEY(chronic kidney disease) OR TITLE-ABS-KEY(chronic kidney failure) OR TITLE-ABS-KEY(CKD) OR TITLE-ABS-KEY(chronic renal failure) OR TITLE-ABS-KEY(chronic renal disease) OR TITLE-ABS-KEY(chronic kidney insufficiency) OR TITLE-ABS-KEY(end stage renal disease) OR TITLE-ABS-KEY(ESRD) OR TITLE-ABS-KEY(kidney function) OR TITLE-ABS-KEY(renal function) OR TITLE-ABS-KEY(kidney dysfunction) OR TITLE-ABS-KEY(renal dysfunction) OR TITLE-ABS-KEY(chronic W/2 kidney W/2 disease) OR TITLE-ABS-KEY(chronic W/2 kidney W/2 failure) AND NOT DBCOLL(medl))

### WEB OF SCIENCE

(((chronic renal insufficiency) OR (chronic kidney disease) OR (chronic kidney failure) OR (CKD) OR (Renal Insufficiency, Chronic) OR (chronic NEAR/2 kidney NEAR/2 disease) OR (chronic NEAR/2 kidney NEAR/2 failure) OR (chronic renal failure) OR (chronic renal disease) OR (chronic kidney insufficiency) OR (end stage renal disease) OR (ESRD) OR (kidney function) OR (renal function) OR (kidney dysfunction) OR (renal dysfunction)) AND ((“Afghanistan”) OR (“Benin”) OR (“Burkina Faso”) OR (“Burundi”) OR (“Central African Republic”) OR (“Chad”) OR (“Comoros”) OR (“Democratic Republic of the Congo”) OR (“Eritrea”) OR (“Ethiopia”) OR (“Gambia”) OR (“Guinea”) OR (“Guinea-Bissau”) OR (“Haiti”) OR (“Democratic People’s Republic of Korea”) OR (“Liberia”) OR (“Madagascar”) OR (“Malawi”) OR (“Mali”) OR (“Mozambique”) OR (“Nepal”) OR (“Niger”) OR (“Rwanda”) OR (“Senegal”) OR (“Sierra Leone”) OR (“Somalia”) OR (“South Sudan”) OR (“Tanzania”) OR (“Togo”) OR (“Uganda”) OR (“Zimbabwe”) OR (“Armenia”) OR (“Bangladesh”) OR (“Bhutan”) OR (“Bolivia”) OR (“Cape Verde”) OR (“Cambodia”) OR (“Cameroon”) OR (“Congo”) OR (“Cote d’Ivoire”) OR (“Djibouti”) OR (“Egypt”) OR (“El Salvador”) OR (“Ghana”) OR (“Guatemala”) OR (“Honduras”) OR (“India”) OR (“Indonesia”) OR (“Kenya”) OR (“Micronesia”) OR (“Kosovo”) OR (“Kyrgyzstan”) OR (“Laos”) OR (“Lesotho”) OR (“Mauritania”) OR (“Moldova”) OR (“Mongolia”) OR (“Morocco”) OR (“Myanmar”) OR (“Nicaragua”) OR (“Nigeria”) OR (“Pakistan”) OR (“Papua New Guinea”) OR (“Philippines”) OR (“Samoa”) OR (“Atlantic Islands”) OR (“Melanesia”) OR (“Sri Lanka”) OR (“Sudan”) OR (“Swaziland”) OR (“Syria”) OR (“Tajikistan”) OR (“Timor-Leste”) OR (“Tonga”) OR (“Tunisia”) OR (“Ukraine”) OR (“Uzbekistan”) OR (“Vanuatu”) OR (“Vietnam”) OR (“Middle East”) OR (“Yemen”) OR (“Zambia”) OR (“Albania”) OR (“Algeria”) OR (“American Samoa”) OR (“Angola”) OR (“Argentina”) OR (“Azerbaijan”) OR (“Republic of Belarus”) OR (“Belize”) OR (“Bosnia and Herzegovina”) OR (“Botswana”) OR (“Brazil”) OR (“Bulgaria”) OR (“China”) OR (“Colombia”) OR (“Costa Rica”) OR (“Cuba”) OR (“Dominica”) OR (“Dominican Republic”) OR (“Equatorial Guinea”) OR (“Ecuador”) OR (“Fiji”) OR (“Gabon”) OR (“Georgia”) OR (“Grenada”) OR (“Guyana”) OR (“Iran”) OR (“Iraq”) OR (“Jamaica”) OR (“Jordan”) OR (“Kazakhstan”) OR (“Lebanon”) OR (“Libya”) OR (“Macedonia (Republic) “) OR (“Malaysia”) OR (“Indian Ocean Islands”) OR (“Mexico”) OR (“Montenegro”) OR (“Namibia”) OR (“Palau”) OR (“Panama”) OR (“Paraguay”) OR (“Peru”) OR (“Russia”) OR (“Serbia”) OR (“South Africa”) OR (“Saint Lucia”) OR (“Saint Vincent and the Grenadines”) OR (“Suriname”) OR (“Thailand”) OR (“Turkey”) OR (“Turkmenistan”) OR (“Venezuela”) OR (developing countr) OR (lowincome countr*) OR (middle-income countr*) OR (low-middle income countr*) OR (upper-middle income countr*)) AND ((risk assessment) OR (risk equation$) OR (risk chart?) OR (risk NEAR/3 tool$) OR (risk assessment function?) OR (risk assessor) OR (risk appraisal$) OR (risk calculation$) OR (risk calculator$) OR (risk factor$ calculation$) OR (risk engine$) OR (risk equation$) OR (risk table$) OR (risk threshold$) OR (risk disc?) OR (risk disk?) OR (risk scoring method?) OR (scoring scheme?) OR (risk scoring system?) OR (risk scal$) OR (risk prediction?) OR (risk algorith$) OR (prediction model$) OR (predictive instrument?) OR (project$ risk?) OR (predictive model?) OR (scoring method$) OR (prediction$ NEAR/3 method$) OR (risk? NEAR/1 assess$) OR (screening) OR (diagnostic test))) NOT ((animal*) OR (“not humans”))

## Notes

**Support:** RMC-L is supported by a Wellcome Trust International Training Fellowship (214185/Z/18/Z).

### Competing Interest Statement

The authors have declared no competing interest.

### Funding Statement

RMC-L is supported by a Wellcome Trust International Training Fellowship (214185/Z/18/Z).

